# Pulmonary thromboembolism in patients after COVID-19 - predictive indicators for correct diagnosis

**DOI:** 10.1101/2021.06.24.21258842

**Authors:** D. Yakova-Hristova, I. Simova, P. Pavlov, M. Hristov, T. Kundurzhiev, N. Dimitrov, T. Vekov

**Affiliations:** Heart and Brain Centre of Excellence, University Hospital, Pleven, Bulgaria; Medical University, Pleven, Bulgaria; Bulgarian Cardiac Institute; Cardiology Hospital Pleven, Bulgaria; Faculty of Public Health, Medical University, Sofia, Bulgaria

## Abstract

**Introduction:** Infection caused by SARS-CoV-2 has been shown to lead to significant procoagulant events, in some cases involving life-threatening pulmonary thromboembolism (PE). Additional conditions complicating the diagnosis are the presence of risk factors for PE in almost all patients with COVID-19, as well as the overlap of the clinical presentation between PE and COVID-19. Materials and Methods: Тherefore we conducted a single-center study at the Heart and Brain Hospital, Pleven in the period December 2020-February 2021. It included 27 consecutively hospitalized patients with recent pneumonia caused by Covid-19 and clinical presentation referring to PE. The cohort was divided into two groups - with and without a definitive diagnosis of PE, proven by CT pulmoangiography. The aim was to find the indicators that predict the presence of PE in patients with acute or Post-acute COVID-19 conditions. Results: Our results show that part of the ECG criteria - S-wave over 1.5 mm in I lead and aVL (p = 0.007), Q-wave in III and aVF (p = 0.020), as well as the D-dimer as quantitative variable (p = 0.025) proved to be independent predictors of PE. The RV/ LV diameter ratios ≥1.0 as well as right ventricular dysfunction showed sensitivity 62.5%, specificity 100%, positive predictive value 100% and negative such 86.4% to verify the PE diagnosisЛ We suggest that the cut-off value of D-dimer of 1032 ng/ml has an optimal sensitivity (Se) of 87.5%, specificity (Sp) 57.9%, positive a predictive value (PPV) 46.7% and negative predictive value (NPV) of 91.7% for the diagnosis of PE (p = 0.021). Conclusion: Against the background of acute and Post-acute COVID-19 conditions ECG and EchoCG criteria remain predictive of PE. We suggest that a higher D-dimer cut-off value should be applied in COVID-19 and post-COVID-19 patients in order to confirm/dismiss the diagnosis PE.

## Introduction

Infection caused by SARS-CoV-2 has been shown to lead to significant procoagulant events, in some cases involving life-threatening pulmonary thromboembolism (PE) [1]. A number of abnormalities have been described in coagulation parameters, which are predictors of poor prognosis in patients with COVID-19 and PE [2]. Due to the lack of large prospective studies, little is known about the pathogenesis underlying PE, caused by COVID-19. [3, 4]

Diagnosis of pulmonary thromboembolism is a challenge in Covid-19-affected patients. Prolonged immobilization and hypercoagulable status are considered to be predisposing factors for the onset of PE. Viral particles provoke a systemic inflammatory response, which in turn leads to a violation of the balance between the procoagulant and anticoagulant state in the body. The purpose of blood clotting is to prevent the loss of blood and immune components. Thrombosis could reduce the entry of microorganisms into the blood. Endothelial dysfunction is blamed as a possible provoker for the development of microthrombosis. As a result of the dysfunction, the endothelial cells lose their basic properties such as vasodilation, antiplatelet activity and fibrinolysis. [1] Additional conditions complicating the diagnosis are the presence of risk factors for PE in almost all patients with COVID-19, as well as the overlap of the clinical presentation between PE and COVID-19. Understanding them would lead to early diagnosis and prevention of potentially fatal complications through the application of timely and appropriate treatment. In the treatment of patients with COVID-19 complicated by PE, the use of systemic fibrinolysis or catheter-targeted therapy should be limited to those strictly indicated. 1, 5] Our aim was to find the indicators that predict the presence of PE in patients with acute or Post-acute COVID-19 conditions.

## Material and methods

### Treatment of COVID-19 patients in Heart and Brain Centre of Excellence, University Hospital, Pleven, Bulgaria

The COVID-19 department of Heart and Brain Centre of Excellence, University Hospital, Pleven, Bulgaria, was opened in November 2020 and in April 2021 it already has 94 beds, 20 of which are intensive with the option for mechanical ventilation. All patients hospitalized with COVID-19 pneumonia received a therapeutic dose of anticoagulant and a prophylactic dose of antiplatelet agent during treatment. At their discharge, an antiplatelet agent was recommended for 1 month, unless there is an indication for longer use of the latter.

### Study group

A single-center study was conducted at the Heart and Brain Hospital, Pleven in the period December 2020-February 2021. It included 27 consecutively hospitalized patients with recent pneumonia caused by Covid-19 and clinical presentation referring to PE. The inclusion criteria were patients ≥18 years of age, with active or experienced Covid-19 pneumonia, with clinical, laboratory and diagnostic criteria for PE and no allergy to iodine-containing contrast agents, who confirmed their participation by written consent. The exclusion criteria were refusal to participate in the test and allergy to contrast. The cohort was divided into two groups - without and with a definitive diagnosis of PE, proven by CT pulmoangiography. During treatment with COVID-19, all patients received a prophylactic dose of anticoagulant and antiplatelet drug. The treatment managment of patients diagnosed with PE was in line with the recommendations of the European Society of Cardiology. Due to the higher risk of bleeding, catheter-targeted thrombolysis with Actilyse was performed according to a protocol. For this purpose, the right femoral vein was used for vascular access and 15-20 ml of Actilyse were injected into the affected branch of the pulmonary artery or bilaterally by means of a pig tail catheter.

### Statistical analyses

Statistical analyzes were performed using statistical software SPSS for Windows version 20.0. Continuous variables were presented as mean±standard deviation (SD). The category variables were presented as a percentage. Comparisons of continuous variables between the two groups were performed with the Mann-Whitney Test. The relationship between diagnosis and categorical variables was evaluated by Fisher’s exact test. Receiver Operating Characteristic Analysis (ROC) was used to determine the diagnostic capabilities of the D-dimer. Sensitivity (Se), Specificity (Sp), positive and negative predictive values (PPV and NPV) were calculated at the cut-off value of D-dimer of 1032 ng/m. A two-tailed p value < 0.05 was considered statistically significant.

### Ethical considerations

All patients signed an informed consent for pulmoangiography and fibrinolysis, and for personal data analysis. The study protocol is in accordance with the Declaration of Helsinki.

## Results

Our results showed that eight patients from the group had PE, and 19 had not evidence of PE. The mean age of the group was 65 years and 18 of the patients were women. Тhe two groups did not differ significantly in age and distribution between the sexes (Table 1).

**Table 1.** Comparison between patients with active or experienced Covid-19 pneumonia without and with definitive diagnosis of PE

| Variable | Patients with active or experienced Covid-19 pneumonia without definitive diagnosis of PE<br>n = 19 | Patients with active or experienced Covid-19 pneumonia with definitive diagnosis of PE<br>n = 8 | p value |
| --- | --- | --- | --- |
| Age (years) (mean ± SD) | 65,37 ± 10,57 | 65,63 ± 11,40 | 0,710 |
| Female (n, %) | 13 (68,4) | 5 (62,5) | 1,000 |
| VTE (n, %) | 1 (5,3) | 1 (12,5) | 0,513 |
| Recent trauma/surgery (n, %) | 1 (5,3) | 1 (12,5) | 0,513 |
| Active neoplastic process (n, %) | 1 (5,3) | 1 (12,5) | 0,513 |
| Previous PE (n, %) | 1 (5,3) | 0 (0,0) | 1,000 |
| Overweight/obesity (n, %) | 14 (73,7) | 7 (87,5) | 0,277 |
| Heart rate ≥ 75/min (n, %) | 13 (68,4) | 8 (100) | 0,153 |
| Symptoms of HF (n, %) | 2 (10,6) | 3 (37,5) | 0,169 |
| SatO2 at admission (%) (mean ± SD) | 90,21 ± 7,40 | 89,00 ± 3,78 | <0,001 |
| ICU stay (n, %) | 4 (21,1) | 1 (12,5) | 1,000 |
| D-dimer ((ng/mL) (mean ± SD) | 1546,00 ± 2082,13 | 6489,75 ± 6127,30 | 0,021 |
| hsCRP (mg/L) (mean ± SD) | 41,48 ± 50,37 | 45,31 ± 41,65 | 0,832 |
| Leu (x 10<sup>9</sup> g/L) (mean ± SD) | 7,75 ± 2,42 | 9,80 ± 3,25 | 0,123 |
| Lym (x 10<sup>9</sup> g/L) (mean ± SD) | 1,25 ± 0,71 | 1,45 ± 1,20 | 0,873 |
| Plt (x 10<sup>9</sup> g/L) (mean ± SD) | 327,95 ± 164,48 | 249,13 ± 80,92 | 0,265 |
| Shortness of breath after experiencing PE (n, %) | 14 (73,7) | 4 (50,0) | 0,375 |
| Edema after experiencing PE (n, %) | 3 (15,8) | 2 (25,0) | 0,616 |
| Cough after experiencing PE (n, %) | 4(21,1) | 2(25,0) | 1,000 |

The two groups did not differ significantly according to demographics and baseline characteristics. In the context of the disease, increased markers of inflammation were observed in all patients. Blood oxygen saturation was markedly lower in the group with definite PE. It is also noteworthy that in most patients shortness of breath and fatigue persist after controlling the disease.

Statistically significant differences in electrocardiographic findings were observed in the two groups. In patients without PE, 18 (94.7%) had no evidence of S-wave greater than 1.5 mm in I, aVL. On the other hand, in the group diagnosed with PE in 3 (37.5%) this ECG criteria was not present, and in 5 (62.5%) it was present (p = 0.004). Similar ratios were found in terms of the presence of Q-wave in III, aVF. In patients without PE, 18 (94.7%) did not have this ECG sign, while it was present in half of the patients with PE(p = 0.017).

Statistically significant differences between the two groups were observed in regard to the indicator - the ratio RV/LV diameters ≥1.0 (p = 0.001). In patients without PE there was none with an increase in the ratio ≥1 in favor of the right ventricle, while in the group of patients with massive form 5 (62.5%) had the ratio RV / LV diameters ≥1.0, and 3 (37, 5%) did not have. The results were demonstrated in patients without PE there was none with right ventricular dysfunction while in the group of patients with massive form 5 (62.5%) had right ventricular dysfunction, and 3 (37, 5%) did not have. (p = 0.001). The RV/ LV diameter ratios ≥1.0 as well as right ventricular dysfunction showed Se 62.5%, Sp 100%, PPV 100% and NPV such 86.4% to verify the PE diagnosis.

D-dimer values differed significantly in the two groups. In patients without PE, the mean D-dimer value was 1546 ng/ml (109-8840), while in those with PE - 6489.75 ng/ml (570-17051) (p = 0.021). For our laboratory, the upper limit of the normal range is 500 ng/ml. As a result of the ROC analysis we found that the cut-off value of D-dimer of 1032 ng/ml (2,064 times above the upper limit of the normal range) has an optimal Se of 87.5%, Sp 57.9%, PPV 46.7% and NPV of 91.7% for the diagnosis of PE (p = 0.021). (Figure 1).

**Figure 1–.**
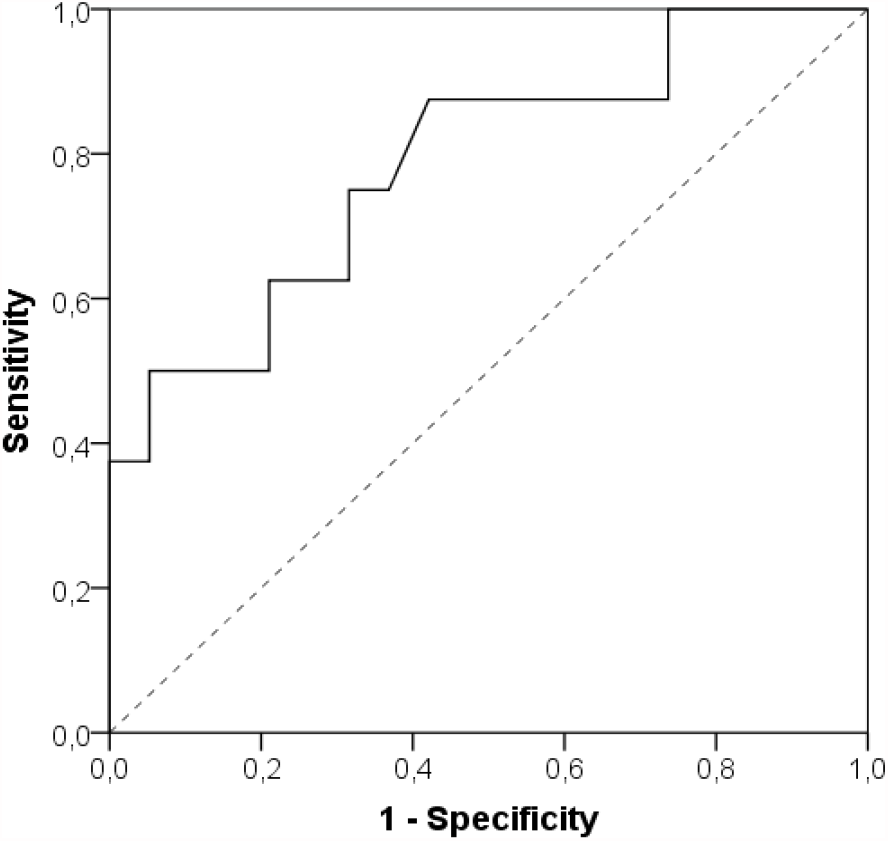
ROC analysis of the cut-off value of D-dimer

Regarding D-dimer as a binary variable (cut-off 1032 ng/ml), we found that in the group without PE, in 11 (57.9%) of patients the D-dimer was ≤ 1032 ng/ml, while in 8 (42.1%) it was > 1032 ng/ml. Of the patients with massive PE, only 1 (12.5%) had a D-dimer ≤ 1032 ng/ml, and the remaining 7 (87.5%) had values > 1032 ng/ml (Fisher’s exact tests, p = 0.043).

When performing binary logistic regression, part of the ECG criteria - S-wave over 1.5 mm in I lead and aVL (p = 0.007), Q-wave in III and aVF (p = 0.020), as well as the D-dimer as quantitative variable (p = 0.025) proved to be independent predictors of PE.

## Discussion

From our results we fоund that ECG and EchoCG criteria remain predictive of PE. A cut-off concentration of D-dimer with optimal Se, Sp, PPV and NPV for diagnosis of PE, is two times higher than the upper limit of normal, with high Se and NPV.

Diagnosis of pulmonary thromboembolism is a challenge in Covid-19-affected patients. The incidence of this complication in hospitalized patients is 1.9-8.9%. Critically ill patients admitted to intensive care units have the highest risk of developing PE, in some cases up to 26.6%. [4, 6, 7] Prolonged immobilization and hypercoagulable status are considered to be predisposing factors for the onset of PE. The hypercoagulable state was confirmed by Hen and co-authors, who demonstrated that higher levels of D-dimer, fibrinogen, prolonged thromboplastin time, prothrombin time, and INR were predictors of poor prognosis in patients affected by SARS-CoV-2. [1, 8] Viral particles provoke a systemic inflammatory response, which in turn leads to a violation of the balance between the procoagulant and anticoagulant state in the body. The immune and coagulation systems are closely linked. The purpose of blood clotting is to prevent the loss of blood and immune components. On the other hand, thrombosis could reduce the entry of microorganisms into the blood. In addition, the constituents of the platelets themselves have antimicrobial activity. [1, 9] Therefore, the body seeks to limit the viral load through thrombosis. Deep venous thrombosis or other sources of non-venous thromboembolism have not been systemically detected in patients with COVID-19 complicated by PE. Endothelial dysfunction is blamed as a possible provoker for the development of microthrombosis. [1, 9, 10] Endothelial cells represent nearly one-third of the cells in the bronchoalveolar tree. As a result of the dysfunction, they lose their basic properties such as vasodilation, antiplatelet activity and fibrinolysis. The endothelial cells themselves have receptors for SARS-CoV-2 - angiotensin-converting enzyme-2 receptors, which facilitate the penetration of viral particles. A number of cytokines released as a result of a systemic inflammatory response lead to endothelial cell apoptosis. [1, 11]

Another predisposing factor for the hypercoagulable state in the body is hypoxia, which increases the viscosity of the blood. A number of risk factors underlie the possibility of developing PE such as age, obesity, family history of PE, heart and respiratory failure, pregnancy, stroke, trauma, surgery, neoplastic diseases. Not to be underestimated is the fact that almost all patients with COVID-19, especially hospitalized, have at least one, and often multiple risk factors for venous thromboembolism. There are predisposing factors for PE in intensive care units such as immobilization, sedation and the use of central venous catheters. [1, 12]

On the one hand, the clinical symptoms in patients with COVID-19, including shortness of breath, fever, cough, are not specific for PE and are explained with the infection. The study of the D-dimer does not give us objective information in this infection and it is debatable to rely on it when considering the possibility of PE. In addition, the use of native computed tomography could not provide specific information for pulmonary thromboembolism. The possibility of developing contrast-induced nephropathy when performing computed tomography pulmoangiography should not be overlooked, especially in patients in shock with severe COVID infection. [4, 13-17] There are a number of logistical problems in computed tomography. These are often microcirculatory occlusions and thrombosis that cannot be represented by this method. Therefore, a stepwise clinical, laboratory and radiological evaluation of patients with COVID should be performed when assessing the likelihood of PE.

A number of data suggest that the prophylactic dose of unfractionated or fractionated heparin improves survival in a number of patients with criteria for sepsis-induced coagulopathy or with very high D-dimer values. [18] Although data are limited, recommendations have also been found for the use of a therapeutic dose of anticoagulants in hospitalized patients with COVID-19. The decision for it must be strictly individual and taken into account in patients with multiple risk factors and those in critical condition. [1, 19, 20]

Algorithms have been developed in which a D-dimer is used in all patients as a triage test to diagnose PE. [21] At low values - 500-1000 ng / ml PE is excluded without the need for CT-pulmoangiography. Above these threshold values it is necessary to implement it. The threshold values of 500-1000 ng/ml in these algorithms have a 100% negative predictive value. [21, 23]

According to Mouhat and colleagues, the threshold above which PE can be most suspected in COVID-patients is 2590 ng/mL. They conducted a retrospective study of 162 patients with severe Covid infection. In this study, this cut-off value had Se 83.3%, Sp 83.8%, PPV 72.9%, NPV 90.5%. Therefore, it can be assumed that this threshold would lead to the omission of nearly 17% of pulmonary emboli. [22, 23] In contrast to the above studies, we found that the cut-off value of D-dimer of 1032 ng / ml (2,064 times above the upper limit of the normal range) has an optimal Se, Sp, PPV and NPV for the diagnosis of PE (p = 0.021). Therefore, all these results suggest the need for further research and validation of a uniform cut-off value.

### Limitations

Our limitations are related to the small selected group of patients, but our work continues in terms of expanding the cohort and presenting more detailed results.

## Conclusion

Our results show that against the background of acute and Post-acute COVID-19 conditions ECG and EchoCG criteria remain predictive of PE. As for the D-dimer values, we found that a cut-off concentration with optimal Se, Sp, PPV and NPV for diagnosis of PE, is two times higher than the upper limit of normal, with high Se and NPV. We suggest that a higher D-dimer cut-off value should be applied in COVID-19 and post-COVID-19 patients in order to confirm /dismiss the diagnosis PE.

